# Setting a course for preventing hepatitis E in low and lower-middle-income countries: A systematic review of burden and risk factors

**DOI:** 10.1101/2020.11.27.20239715

**Authors:** Aybüke Koyuncu, Daniel Mapemba, Iza Ciglenecki, Emily S. Gurley, Andrew S. Azman

**Affiliations:** Independent Researcher, Atlanta, Georgia; South African Field Epidemiology Training Program, National Institute for Communicable Diseases, Division of National Health Laboratory Services, Johannesburg, South Africa; Médecins Sans Frontières, Geneva, Switzerland; Department of Epidemiology, Johns Hopkins Bloomberg School of Public Health, Baltimore, Maryland

**Keywords:** hepatitis E, hepatitis E virus (HEV), seroprevalence, outbreaks, risk factors

## Abstract

**Background:** Hepatitis E virus is responsible for outbreaks of acute jaundice in Africa and Asia, many of which occur among displaced people or in crisis settings. While an efficacious vaccine for HEV has been developed, we lack key epidemiologic data needed to understand how best to use the vaccine for hepatitis E control in endemic countries.

**Methods:** We conducted a systematic review of articles published on hepatitis E in low and lower-middle-income countries (LMIC) in Africa and Asia. We searched PubMed, Scopus, and Embase databases to identify articles with data on anti-HEV IgG seroprevalence, outbreaks of HEV, or risk factors for HEV infection, disease, or death, and all relevant data were extracted. Using these data we describe the evidence around temporal and geographical distribution of HEV transmission and burden. We estimated pooled age-specific seroprevalence and assessed the consistency in risk factor estimates.

**Results:** We extracted data from 148 studies. Studies assessing anti-HEV IgG antibodies used 18 different commercial assays. Most cases of hepatitis E during outbreaks were not confirmed. Risk factor data suggested an increased likelihood of current or recent HEV infection and disease associated with fecal-oral transmission of HEV, as well as exposures to blood and animals.

**Conclusion:** Heterogeneity in diagnostic assays used and exposure and outcome assessment methods hinder public health efforts to quantify burden of disease and evaluate interventions over time and space. Prevention tools such as vaccines are available, but require a unified global strategy for hepatitis E control to justify widespread use.

## Introduction

Hepatitis E virus (HEV) is a single-stranded RNA virus which causes over 3 million cases of symptomatic hepatitis each year (1). The global burden of HEV is predominantly attributable to genotypes 1 and 2, which are associated with sporadically occurring cases as well as large protracted outbreaks in low and middle income countries where access to clean water and sanitation is limited (1,2). HEV outbreaks disproportionately affect pregnant women, for whom the mortality rate due to hepatitis E has been reported to be as high as 25% (3).

While outbreaks of HEV genotypes 1 and 2 in low and lower-middle-income countries (LMIC) are caused by fecal contamination of drinking water, transmission of other HEV genotypes, predominantly 3 and 4, can cause acute sporadic cases of hepatitis. Genotypes 3 and 4 are non-epidemic, are known to circulate in animal reservoirs, and can be spread through zoonotic transmission, consumption of raw or undercooked meat, blood transfusions, and organ donation (2–4). Although acute sporadic cases of HEV in high-income countries are typically associated with genotypes 3 and 4, the burden of these genotypes in LMIC remains unclear (2).

Access to clean water and sanitation could significantly reduce the transmission of HEV in LMIC, but household-level water and sanitation interventions often require behavioral change and have had mixed results in outbreak control (5). Outbreaks of HEV, many of which occur among displaced people or in crisis settings, are notoriously difficult to control in part due to the high proportion of asymptomatic and mildly symptomatic infections and limited options for clinical management of severe cases (3). One potential intervention strategy to control HEV transmission is a recombinant vaccine, HEV239 (Hecolin®; Innovax, Xiamen, China), which has been shown to be safe and highly efficacious in preventing HEV infection due to genotype 4, with pan-genotype protection expected (6). The World Health Organization (WHO) has not recommended the routine use of the vaccine in HEV endemic countries, citing a critical lack of epidemiologic data on the incidence of disease in the general population and in special subpopulations (7). In the absence of these data Hecolin is currently only recommended by WHO for consideration during HEV outbreaks, although at the time of writing this it has not been used in any outbreaks (7).

Burden-of-disease data, combined with information on key risk factors for HEV infection and disease can spur the use of existing prevention tools that may be capable of substantially reducing morbidity and mortality due to HEV in LMIC. A lack of data on the burden and transmission patterns of HEV precludes investments into a global strategy for control for hepatitis E (8). Given the clear pathway for WHO prequalification of the existing vaccine (9), a better understanding of the epidemiology of hepatitis E viruses is needed to set the agenda for global strategy and identify best practices to measure impact. Here we aim to review data on the epidemiology of hepatitis E in low and lower-middle-income countries in Africa and Asia through systematic reviews of three domains of evidence, first through a review of seroprevalence studies, second a review of reported outbreak size and geography, and finally a review of risk factors for infection, disease and death.

## Methods

### Search Strategy

We searched PubMed, Scopus, and Embase to identify all studies with ‘hepatitis E’ in the article title that were published prior to November 2018, regardless of language. We uploaded search results to covidence (www.covidence.org), which we used to manage the subsequent steps of the review.

### Study Selection

For both stages of study selection (abstract review, full text review) two independent reviewers screened each document for eligibility. All discrepancies between reviewers were reviewed by a third reviewer, who discussed with the original reviewers and decided upon the correct classification.

Studies were included if they satisfied the following inclusion criteria: (i) study in a low-income or lower-middle-income country in Africa or Asia according to 2019 World Bank country classifications (10); and (ii) includes serosurvey data (anti-HEV IgG only, IgG and/or IgM) in populations without known liver disease; or (iii) includes an outbreak description of HEV genotype 1, 2, or unidentified genotype; or (iv) includes effect size estimates (e.g. odds ratio, risk ratio) for potential risk factors for current or recent infection (anti-HEV IgM or RNA), disease, or death.

Studies were excluded if they met any of the following criteria: (i) abstract or full-text not available; (ii) abstract or full-text not in French, English or Spanish; (iii) non-human study of HEV; (iv) diagnostic, viral genetics, or immunologic study; (v) case series with no controls for any outcome; (vi) commentary, perspective, or review article; (vii) studies with duplicate data.

### Data Extraction

A data extraction form developed by the authors was used to extract data from the full-text of all included studies. A plot digitizer (https://apps.automeris.io/wpd/) was used if data were only presented graphically. To gauge the consistency of the data extraction and to identify common errors, 10% of all studies in the data abstraction phase were extracted independently by two reviewers, and any discrepancies in data extracted were resolved by discussion and consensus. We used lessons from discrepancies in extracted data to adjust previous extractions and to ensure that future extractions did not have those errors.

### Seroprevalence Studies

For studies reporting seroprevalence data, we extracted population type, lower and upper bounds for population age, diagnostic assay, test isotype (anti-HEV IgG or IgG+IgM), and the total number of samples tested and number of positive/negative samples. Within each study, data were extracted separately for each age stratum and unique population type: general population, blood donors, patients attending a hospital or clinic for non-liver related, non-pregnancy related issues, pregnant women, occupational groups exposed to animals (e.g. butchers, pig farmers), outbreak residents (i.e. individuals identified in areas with an ongoing HEV outbreak) or other. Refugee and internally displaced populations that were not experiencing outbreaks were categorized as “other”. If studies reported seroprevalence data from the same population using different assays including Wantai assays (Wantai BioPharm, Beijing, China; considered the most reliable commercially available assay at the time of writing), we only extracted seroprevalence data based on Wantai assays (11).

### Outbreak Descriptions

For studies describing outbreaks, we extracted the date of the first and last cases reported in the outbreak, whether the outbreak occurred in a refugee or displaced persons camp, HEV genotype(s) identified, suspected/confirmed case definitions and laboratory confirmation methods, number of suspected cases with confirmatory testing, average age (mean or median) of cases, number of suspected/confirmed cases, and number of suspected/confirmed deaths. Data on the number of suspected/confirmed cases and deaths were extracted within stratum of where cases were identified, namely: at health facilities, in communities, in institutions (e.g. prisons, army barracks), and not reported. The number of suspected/confirmed cases and deaths was extracted for the general population and also separately for pregnant/postpartum women and children under the age of 5. If multiple studies described the same outbreak (overlapping outbreak dates in the same geographic area and population) the most comprehensive outbreak report was extracted. Studies describing the same outbreak were combined only if they presented unique data (e.g. data on different subpopulations, number of deaths, etc.).

### Risk Factors

For studies including risk factors for infection, disease, or death, we extracted outcome definition and confirmation method, estimator of association (e.g. odds ratio, risk ratio), effect size, 95% confidence interval, reference category for the estimator, and whether the estimator was adjusted for any covariates using regression analysis or matching. If both odds ratios and risk ratios were presented for the same risk factor, we extracted odds ratios. If risk factor data were reported for different case definitions of infection/disease within the same study, we extracted the most conservative case definition. We categorized each risk factor into one of the following groups: water, sanitation, hygiene, food and drink (non-water), exposure to blood or blood products, exposure to animals, age, sex, or other. Current and recent HEV infection and disease was defined in three categories: serologic evidence of anti-HEV IgM or RNA with no accompanying symptoms (“confirmed infection”), symptoms consistent with hepatitis (e.g. jaundice, elevated liver enzymes) without any serologic testing (“suspected disease”), or symptoms consistent with hepatitis with confirmation by IgM or HEV RNA (“confirmed disease”).

### Data Analysis

We examined trends in the number of seroprevalence studies conducted over time, as well as variability in seroprevalence by age, population type, and diagnostic assay. Age-specific seroprevalence was calculated using the median age of tested individuals for each seroprevalence estimate. Seroprevalence estimates with no data on population age were excluded from analysis of seroprevalence by age.

We estimated outbreak duration using the difference between the date of the first and last cases reported in each outbreak description. The case fatality risk (CFR) was calculated separately among suspected cases and confirmed cases, and was defined as the number of suspected/confirmed deaths divided by the number of suspected/confirmed cases.

We categorized risk factors based on the general consensus of the literature on factors that increase HEV infection/disease/death risk and those that are protective. Risk factors lacking a hypothesized direction of effect (e.g. ethnicity) or with an unspecified reference group were excluded from further analysis. Household water sources and type of sanitation facilities were categorized as improved or unimproved according to 2019 World Health Organization/United Nations Children’s Fund standard categories (12). A summary of risk factors and their corresponding exposures hypothesized to increase the likelihood of HEV infection/disease/death is provided in the Appendix (Supplemental Table 1). If studies reported unadjusted and adjusted estimates we included the adjusted estimates. We examined the proportion of risk factors that followed the hypothesized direction of effect (e.g. effect estimate >1 for risk factors hypothesized to increase risk) and the proportion that were statistically significant based on 95% confidence intervals or a p-value < 0.05.

## Results

After the exclusion of duplicates, our search identified 4,928 studies for abstract screening, of which 666 met the criteria for full-text review. We identified 148 studies that satisfied the inclusion criteria after full text review (Figure 1).

**Figure 1.**
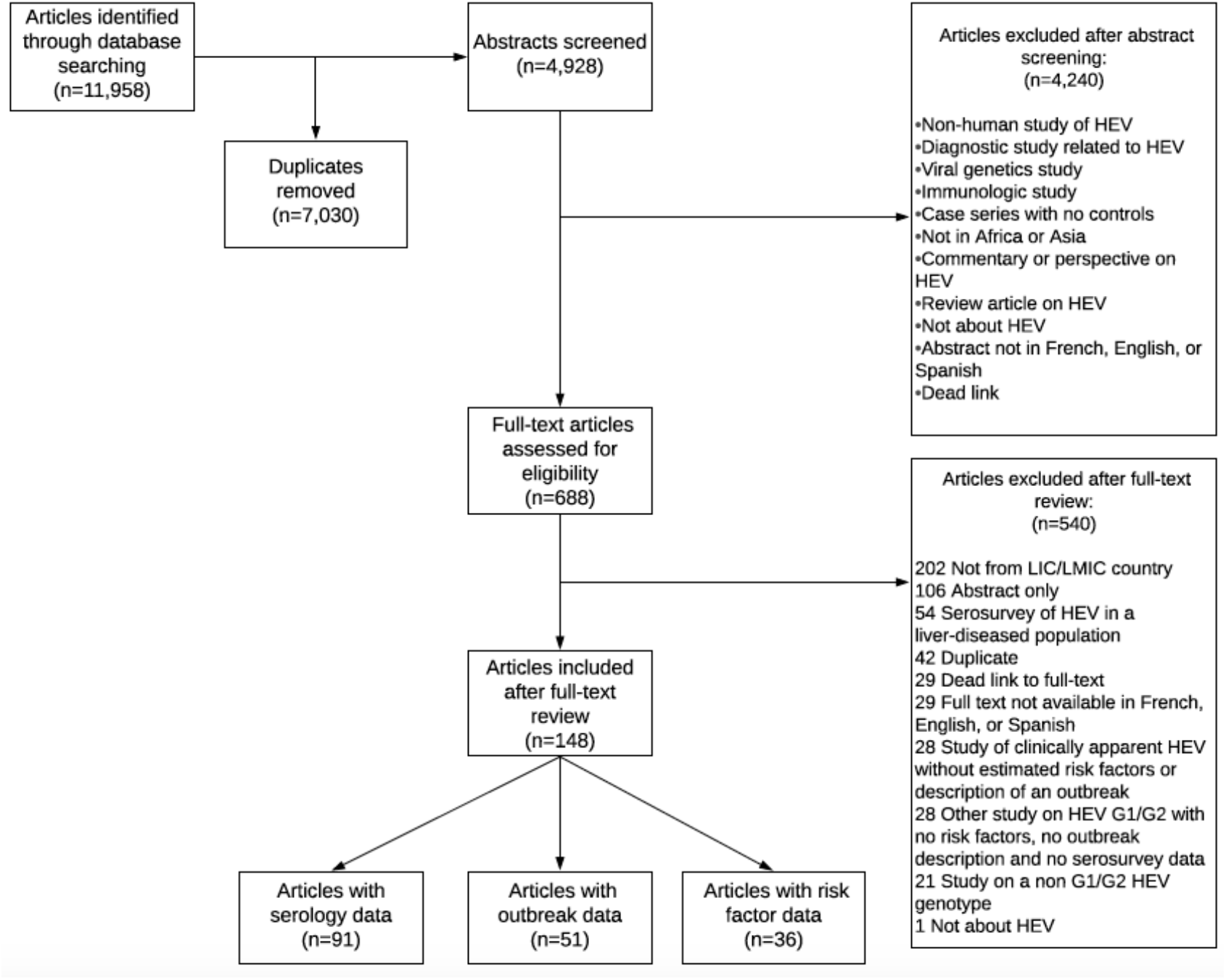
Flow diagram for study screening and selection

### Seroprevalence

We identified 91 studies with seroprevalence data (13–103) from 29 countries, with serum collected between 1978 and 2017 (Figure 2).

**Figure 2.**
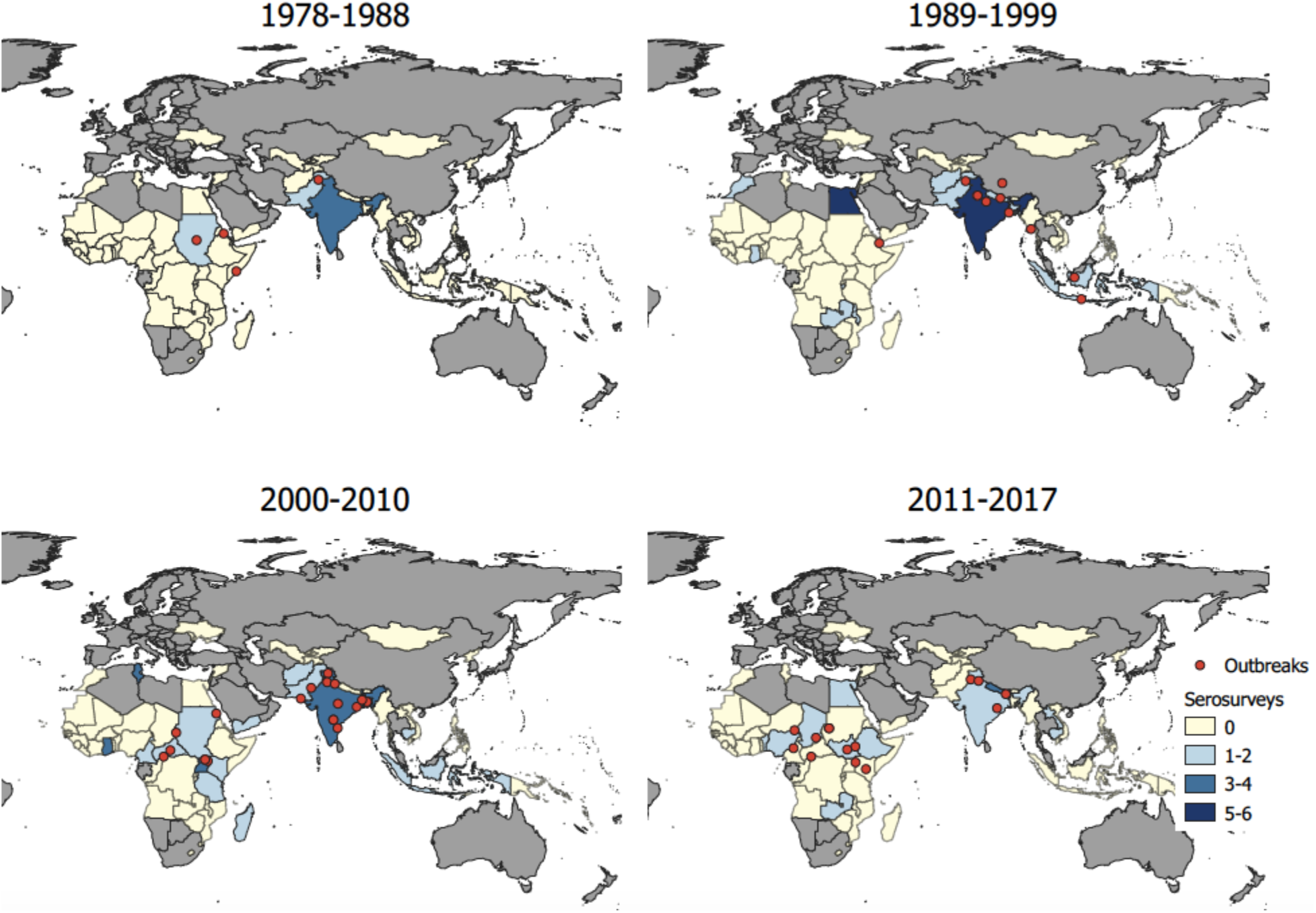
Studies including HEV outbreak and serosurvey data over time, 1978-2017. Dots represent outbreak locations (centroid of lowest geographic unit of the outbreak identified) and colored polygons represent the number of serosurveys conducted in each country. Excludes 18 seroprevalence studies with unspecified study year.

Among studies that reported assay manufacturer (85%; 77/91), anti-HEV IgG antibodies were assessed using 18 different commercial assays as well as in-house assays (e.g. Walter Reed Army Institute of Research in-house assay). The most common assays used to assess seroprevalence were in-house assays (21%) (14,34,44,47,51,53,56,60,61,70,72,79,84,91,97,100), Wantai (18%; Wantai BioPharm, Beijing, China) (20–23,31,40,46,48,54,59,63,65,76,77), and Abbott (14%; (Abbott Diagnostika, Germany) (15,18,28,32,43,80,82,83,87,93,94) with the Wantai assay becoming the most commonly used assay in studies conducted in the last 5 years of the review (2013-2017).

HEV seropositivity, in general, increased with age among all population groups except occupational groups exposed to animals (Figure 3).

**Figure 3.**
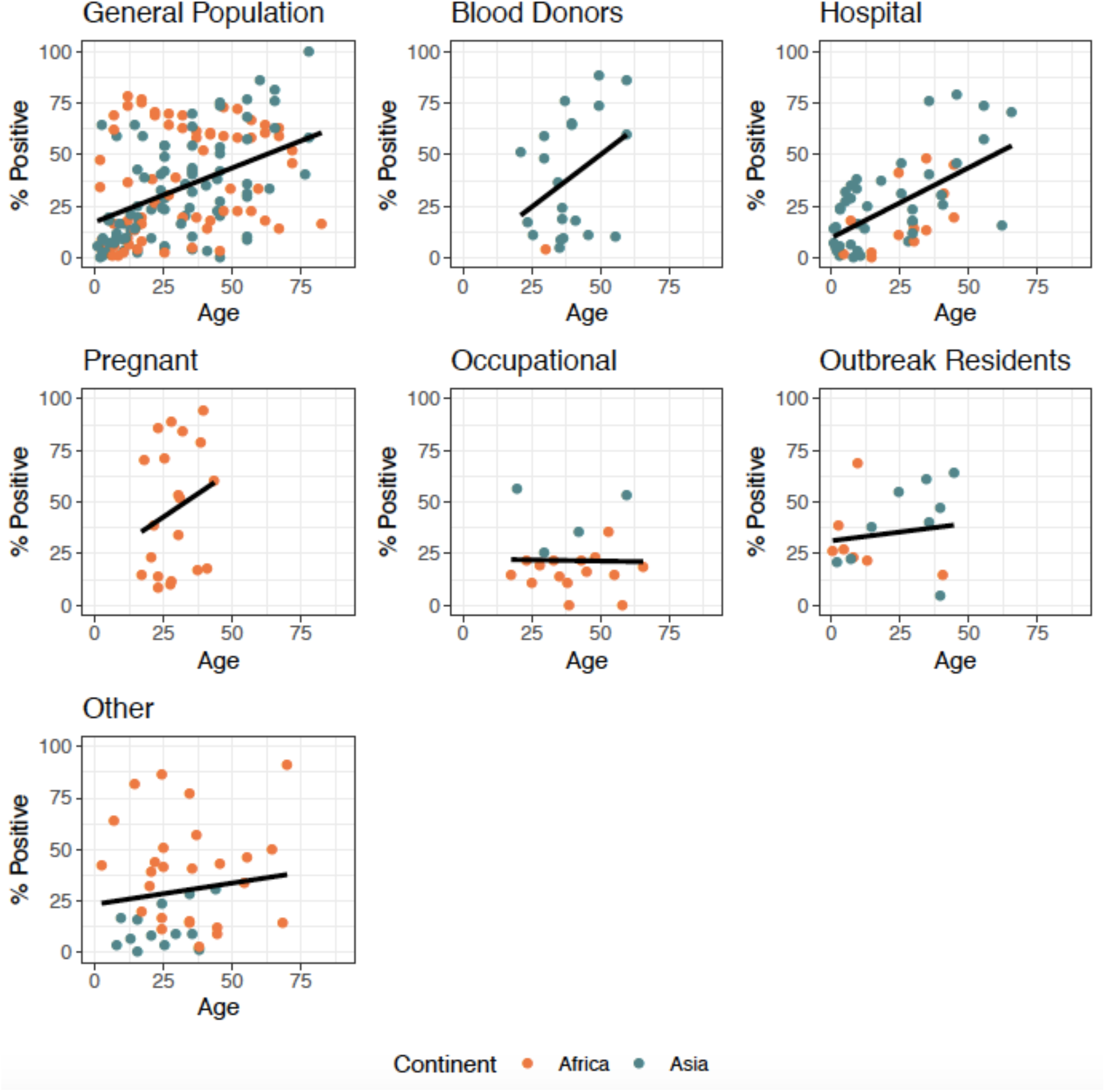
Age seroprevalence curves by varying population types in 29 countries, 1978-2017. Excludes seroprevalence estimates with no specified age bounds and 1 study with unspecified location. Population types include general population, blood donors, patients attending a hospital or clinic for non-liver related, non-pregnancy related issues (hospital), pregnant women, occupational groups exposed to animals (occupational), outbreak residents, or other. Black line represents the best fit linear trend between age and seropositivity.

Age-specific seroprevalence estimates, excluding occupational groups and outbreak residents, increased by age with all assays used, except Abbott (Supplemental Figure 1).

### Outbreaks

We identified 51 outbreak reports (30,50,78,82,85,93,104–107,107–148) describing 49 completed or on-going outbreaks occurring in 18 countries between 1988 and 2017. Among outbreaks with reported locations (48/49), outbreaks were identified in Africa (43%) and Asia (57%), with over half of outbreaks reported from 3 countries: India (35%) (30,106,107,109,110,114,117,121,125,128,136–138,141,145,147,148), Sudan (10%) (119,133,140,143,144), and Uganda (8%) (78,85,108,126,129). Reported outbreak durations ranged from 28 days (Yangon, Myanmar) (142) to 3.2 years (Karamoja region, Uganda) (126) (Median: 166 days; IQR: 91-303). At least one large-scale outbreak with over 5,000 suspected cases occurred in every decade since 1988, and outbreaks in the last 15 years (2004-2017) had a median of 1152 suspected cases reported (IQR: 252-2362). Among all outbreaks, 12% occurred in refugee camp/informal settlements (N=6) (50,78,111,119,123,123), with 31% of all suspected cases (24,341/77,372 suspected cases) identified in our review occurring in these settings.

Confirmatory serologic testing (i.e., IgM) was conducted for 10% of all suspected cases reported in outbreaks (7,956/77,372), including 15 small outbreaks (<500 suspected cases) in which 100% of all suspected cases had confirmatory serologic testing (30,82,93,113,118,121,122,125,129,134,135,140,142–144). Among outbreaks where at least some cases were laboratory confirmed (N=47), 94% of confirmed case definitions included serologic confirmation of HEV IgM and 21% included HEV RNA. Overall, 43% of all suspected cases were identified in health facilities, 42% were identified within the community, 1% were identified in institutions (e.g. military training camps, prisons), and 13% were identified in unknown locations.

Among studies reporting data on deaths (34/49), the CFR ranged from 0-28% among suspected cases (mean: 2; SD: 6) and 0-28% among confirmed cases (mean: 3; SD: 7). Fewer outbreak reports had available data on deaths among pregnant women (20/48), and the CFR among pregnant women ranged from 0-33% among suspected cases (mean: 11; SD: 10) and 0-65% among confirmed cases (mean: 28; SD: 24) (50,78,82,104,106,108,112–114,117,119,120,123,124,126–129,140,146). The highest CFR (65%; 15 fatalities among 23 confirmed pregnant cases) was reported among pregnant women in an outbreak in Napak district, Uganda which had over 1,300 confirmed cases of HEV in the overall population in 2013-2014 (108).

### Risk Factors

Data on risk factors for HEV infection (defined by IgM antibodies or RNA), disease, and death were identified in 36 studies (17,21,26,30,34,46,63,68,74,85,91,92,102,104,108–110,112,116,129,138,139,145–158) from 13 countries between 1990 and 2017. One study only reported risk factors with an unknown hypothesized direction of effect and was excluded from further analysis (159). We identified only 2 studies (112,154) that examined risk factors for death and decided not to further summarize these limited data. Risk factors were predominantly for confirmed disease (65%; 182/279), confirmed infection (25%; 70/279), followed by suspected disease (8%; 23/279), and a mixture of suspected/confirmed disease (1%; 4/279). Studies investigating risk factors occurred in Africa (41%) and Asia (59%), and 33% of all individual risk factor estimates came from studies conducted in Bangladesh (112,139,151,158). Most risk factor estimates included 95% confidence intervals (97%; 270/279) and were not adjusted for other covariates (74%; 206/279). The most common risk factors examined were those categorized as “other” (24%; 68/279) and water (16%; 46/279). 43% of risk factors categorized as “other” (29/68) were related to demographics and socioeconomic status (e.g. rural residence, household size, etc.; Supplementary Table 1). Risk factors related to water included household sources of water for drinking, cooking, or bathing, and water treatment and storage methods. 46% of all studies investigating risk factors (16/35) were from HEV outbreaks (30,85,104,108–110,112,116,119,129,138,139,145–148).

Based on the direction of effect estimates, 67% of effect estimates for water suggested increased risk of HEV with increased exposure to unimproved water sources and 33% of estimates showed the opposite trend or no effect (effect size = 0) (Figure 4). Effect estimates for sanitation and hygiene were similarly heterogeneous: 83% of sanitation-related estimates suggested increased risk with exposure to unimproved sanitation, and 71% of hygiene-related effect estimates suggested increased risk due to poor hygiene practices (e.g., never washing hands after defecating, etc.). Among estimates reported with a measure of statistical significance (76/79), a higher proportion of estimates for sanitation were statistically significant (33%) compared to estimates for water (21%) or hygiene (29%).

**Figure 4.**
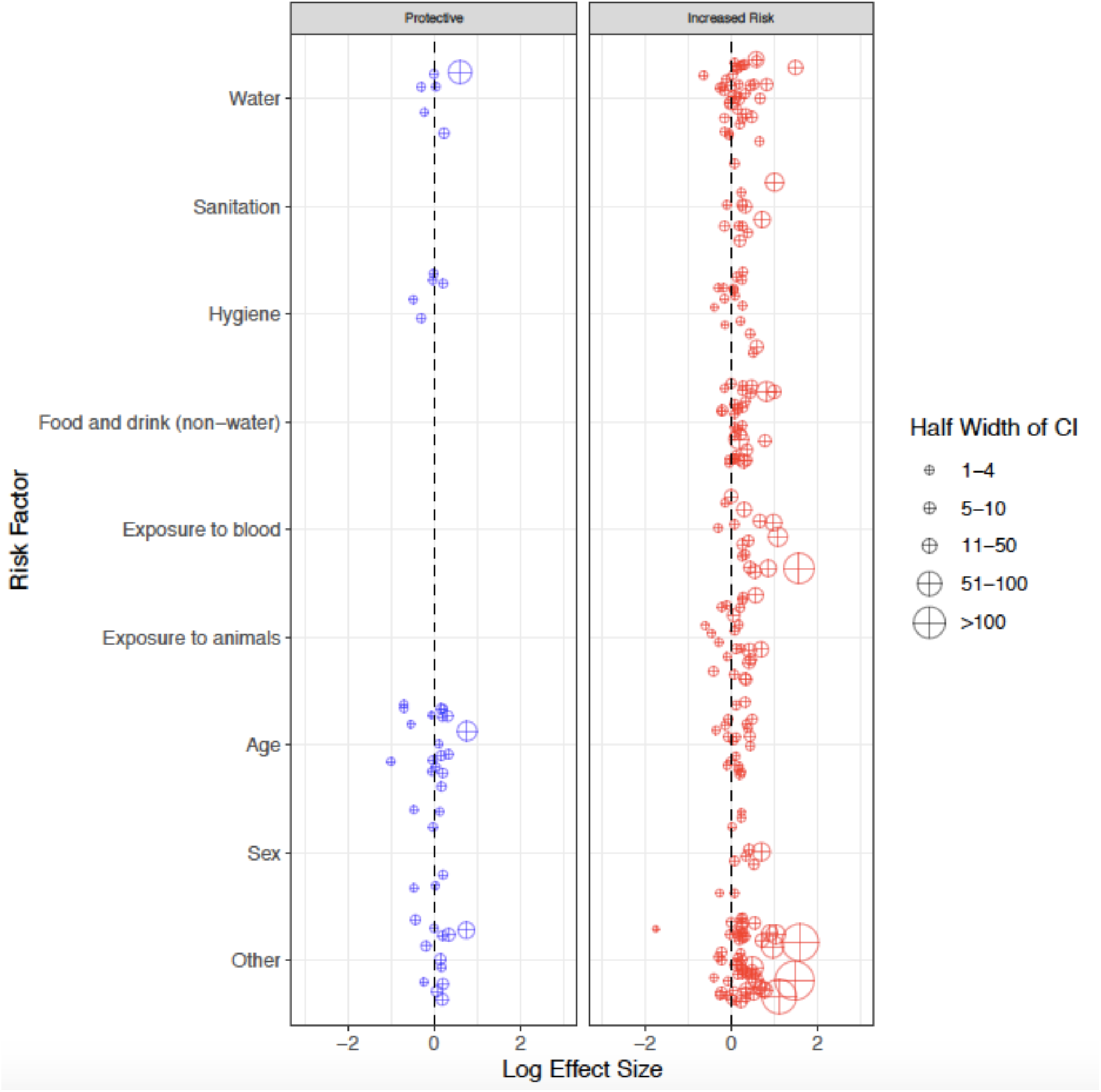
Risk factors for HEV infection or disease in 13 countries, 1990-2017. Risk factors were divided *a priori* into those hypothesized to decrease (blue) or increase (red) the likelihood of HEV infection or disease based on existing literature and the reference group for each estimator (Supplemental Table 1). CI: 95% Confidence Interval.

Excludes 9 risk factor estimates not reported with corresponding 95% confidence intervals.

Increased risk of HEV infection/disease (effect size >1) was identified in a majority of effect estimates related to exposures to food and drink (82% of estimates), blood (81%), and animals (72%). Food and drink exposures (N=33), were predominantly assessed with questions related to consumption of meat (42%) and juices/sodas (24%). The majority of assessed exposures to blood (N=16) were related to blood transfusions (38%) and recent injections (25%). The majority of assessed exposures to animals (N=25) were related to the presence of animals (pets or varmints) in compounds/homes (32%), occupational exposures (24%), any contact with animals (16%), and livestock ownership (16%). Only 21% (7/33) of effect estimates related to food/drink exposures were statistically significant, while 38% (6/16) of estimates for blood exposures and 20% (5/25) of estimates for animal exposures were significant.

The directionality of effect estimates was consistent with the hypothesis that older age increases risk of HEV infection/disease in 52% of estimates for confirmed disease and 54% of estimates for confirmed infection, as would be expected by chance. Effect estimates related to age were statistically significant for 40% (10/25) of estimates for confirmed disease and 8% (1/12) of estimates for confirmed infection. 82% (9/11) of estimates that were statistically significant suggested increased risk for confirmed HEV disease/infection with older age.

In 72% of estimates (13/18) males had a higher risk of infection/disease than females. Among estimates reported with a measure of statistical significance (17/18), 41% were statistically significant.

## Discussion

Through a synthesis of published literature we highlight the persistent burden of hepatitis E genotypes 1 and 2 in LMICs and suggest that zoonotic and blood borne transmission may be contributing to this burden in addition to fecal-oral transmission. Although studies on HEV burden have expanded geographically over time, existing data are insufficient to identify temporal or geographic trends in risk. Similar to other reviews, we note challenges in interpreting and comparing data from different studies due to variability in diagnostic assays used, definitions of suspected and confirmed cases of HEV (160–162), and lack of standardized age groups used in analysis. Unified guidance and best practices for measuring disease burden and risk factors are urgently needed to generate evidence needed for developing a global strategy for hepatitis E control.

Given that most cases of hepatitis E are mild or asymptomatic and the lack of routine testing for hepatitis E among acute jaundice cases in LMICs, our knowledge on HEV burden of disease is largely limited to special studies and outbreak reports (163). Gaps in burden of disease data identified by WHO’s Strategic Advisory Group of Experts on Immunization (SAGE) in 2015 as barriers to widespread introduction of hepatitis E vaccine remain largely unaddressed (7). A lack of data on the incidence of disease in the general population prevents the necessary investments in hepatitis E surveillance infrastructure, which cyclically limits our ability to quantify disease burden and advocate for expanded use of the existing vaccine. Notably, the SARS-CoV-2 pandemic provides an opportunity to break this cycle due to the increased number of representative population-based surveys that have been conducted (164). Leveraging remnant samples from these studies could provide a cost-effective way of greatly increasing our understanding of the global burden of hepatitis E (165).

Our findings emphasize how a reliance on data from outbreak reports can obscure potentially important HEV transmission pathways. Trends suggestive of increased risk for infection and disease due to exposures to animals and consumption of meat suggest that HEV genotypes 3 and 4 may be responsible for some of the hepatitis E burden. Our findings suggest that sporadic hepatitis cases in LMIC may also be due to blood borne transmission of HEV, which is most likely to be caused by the predominant circulating genotype within each population (151). However, our ability to draw conclusions about the role of zoonotic and blood-borne transmission is limited given that only 27% of studies with data on risk factors adjusted for potentially confounding variables such as socioeconomic status. Our ability to identify key risk factors is further limited by the fact that most ‘controls’ in case-control studies were recruited on the basis of not having hepatitis E disease with the assumption that they were still at-risk. Some of these controls were almost certainly previously exposed to HEV, which would lead to reduced power, and in some cases bias, in estimates of risk factors (166). A better understanding of the frequency of HEV infection due to transmission routes such as zoonotic transmission and blood-borne transmission in endemic countries may justify potentially life-saving interventions such as blood donor screening for HEV.

WHO has recommended the vaccine be considered as a strategy to prevent and mitigate outbreaks, particularly among high-risk groups such as pregnant women (7). The current vaccine’s three dose schedule given across 6 months introduces logistical constraints to rapidly deploying vaccines in outbreaks (167). While reduced dose regimens may confer protection against HEV (6), the multi-year protracted outbreaks identified in our review suggest the potential for even a three dose regimen to substantially prevent morbidity and mortality among the world’s most vulnerable populations. While we note that CFRs may be overestimated in studies relying on health facilities for case detection, the average case fatality risk among confirmed pregnant cases in outbreaks in this review was 9.3 times higher than confirmed cases in the general population. Field studies are urgently needed to evaluate the safety and effectiveness of the existing hepatitis E vaccine subpopulations such as refugees and pregnant women who are disproportionately affected by the burden of HEV.

This review has several limitations. We did not conduct any formal risk of bias assessments, and did not exclude any studies on the basis of sample size. Trends in risk factor data may have been under or overestimated due to the inclusion of low quality estimates. We underestimated the number of HEV outbreaks in LMIC due to limited publishing of outbreaks in peer-reviewed journals and our exclusion of less formal outbreak reports, such as the WHO Outbreaks and Emergencies bulletin. Outbreak reports were often based on research investigations of outbreak etiology rather than comprehensive assessments of outbreak size, leading to an underestimation of the burden of outbreaks. Age-specific seroprevalence curves stratified by population type were generated using data from diverse diagnostic assays which have varying sensitivity and specificity, and therefore may not represent true age-specific patterns. Finally, our ability to draw conclusions from risk factor data is limited given our inclusion of unadjusted and adjusted estimators collected from varying population types. Nevertheless, we identify key trends in the force of infection of HEV and risk factors for transmission that warrant further investigation in future studies.

Addressing outstanding gaps in our understanding of the epidemiology of HEV in LMICs is critical to justify the funds needed for expanded use of the vaccine following pre-qualification. Standardized guidelines defining best practices for study design and exposure and outcome measurement in hepatitis E research are needed to identify key areas with elevated incidence of HEV infection where the vaccine may be a cost-effective tool to prevent avoidable morbidity and mortality. Guidelines are also needed to define a minimum set of risk factors for which data should be collected in all HEV outbreak investigations and risk factor studies, including: sociodemographic variables (e.g. SES, age, sex), WASH, and exposures to animals and blood. Broad control of hepatitis E with existing tools and strategies is possible, but requires a unified global strategy and the inclusion of hepatitis E in the agendas of researchers, public health authorities, and funders.

## Data Availability

The data that support the findings of this systematic review are openly available at https://github.com/akoyuncu4/hepatitis-E-in-LMIC-a-systematic-review.git.

