## Supplemental Table 1 for "Setting a course for preventing hepatitis E in low and lower-middle-income countries: A systematic review of burden and risk factors"

Supplemental Table 1. Risk factors for hepatitis E virus (HEV) infection, disease, or death. Household water sources and type of sanitation facilities were categorized as improved or unimproved according to 2019 World Health Organization/United Nations Children’s Fund standard categories for improved/unimproved water and sanitation

| **Risk Factor Type** | **Exposures hypothesized to increase the likelihood of HEV infection, disease, or death** |
| --- | --- |
| Water | -Unimproved household water sources  -Consumption of water from unimproved sources during a predetermined time frame  -Storage of water in a reservoir or wide mouthed container |
| Sanitation | -Unimproved household sanitation facility |
| Hygiene | -Poor hygiene behaviors |
| Animal related | -Contact with animals  -Occupations with possible contact with animals (e.g. farming)  -Presence of animals in the home or on one's property  -Use of dung (e.g. home construction, fuel) |
| Blood | -Receipt of blood transfusion, injections, or intravenous drugs  -Reporting cuts with blood to blood contact  -Touching other people's blood  -Shaving in a barbershop |
| Food and drink (non-water) | -Consumption of a specified food or non-water drink during a predetermined time frame |
| Age | -Older age |
| Sex | -Male sex/gender |
| Other demographics/socioeconomic status (SES) | -Less education  -Lower household income and/or expenditures  -Lower quality (e.g. expense, durability) household construction materials  -More people in the household  -More children in the household  -Higher ratio of household occupants to sleeping rooms in household  -Married  -Rural residence |
