## Supplemental Figure 1 for "Setting a course for preventing hepatitis E in low and lower-middle-income countries: A systematic review of burden and risk factors"

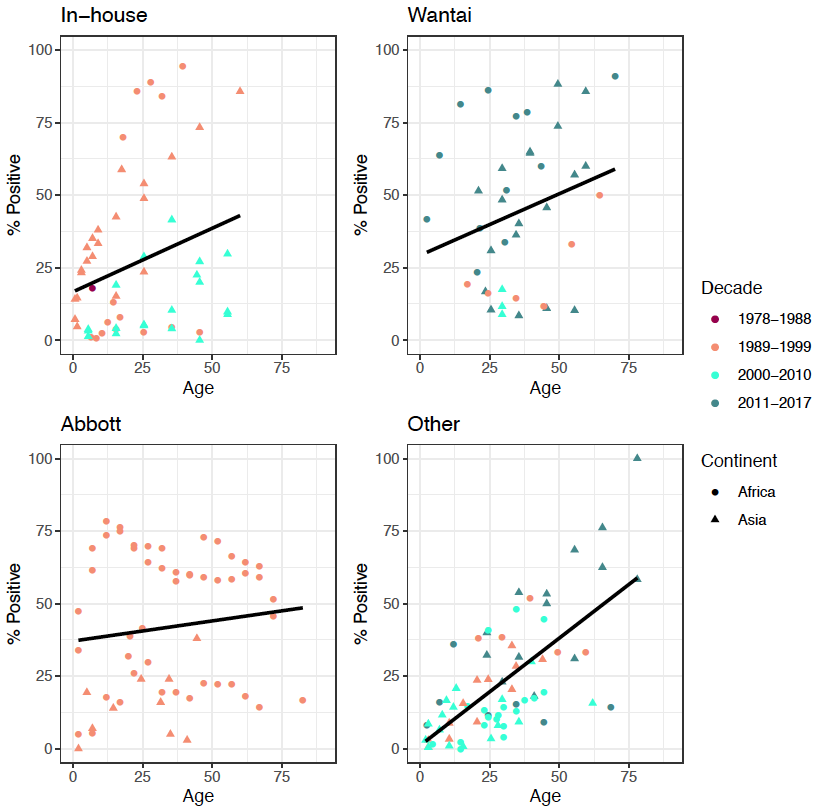

Supplemental Figure 1. Age seroprevalence curves by assay in 29 countries, 1978-2017. Excludes seroprevalence estimates from occupational groups exposed to animals and residents of outbreak affected areas, seroprevalence estimates with no specified age bounds, 6 studies with unspecified study year, and 1 study with unspecified study location. Black line represents the best fit linear trend between age and seropositivity. NIH: National Institutes of Health.
